# Putting Bicarbonate on the spot. Implication for theratyping in Cystic Fibrosis

**DOI:** 10.1101/2023.06.24.23291859

**Authors:** Miroslaw Zajac, Agathe Lepissier, Elise Dréano, Benoit Chevalier, Aurélie Hatton, Mairead Kelly, Daniela Guidone, Gabrielle Planelles, Aleksander Edelman, Emmanuelle Girodon, Alexandre Hinzpeter, Gilles Crambert, Iwona Pranke, Luis J.V. Galietta, Isabelle Sermet-Gaudelus

## Abstract

Cystic fibrosis (CF) is caused by defective Cystic Fibrosis Transmembrane Conductance Regulator (CFTR) protein. CFTR controls chloride (Cl^-^) and bicarbonate (HCO_3_^-^) transport into the Airway Surface Liquid (ASL).We investigated the impact of F508del-CFTR correction on HCO_3_^-^ secretion by studying transepithelial HCO_3_^-^ fluxes.

HCO_3_^-^ secretion was measured by pH-stat techniquein primary human respiratory epithelial cells from healthy subjects (WT) and people with CF (pwCF)carrying at least oneF508del variant.Its changes after CFTR modulation by the triple combination VX445/661/770 and in the context of TNF-α+IL-17 induced inflammation were related to ASL pH and transcriptionnal levels of CFTRand other HCO_3_^-^ transporters ofairway epithelia such asSLC26A4 (Pendrin), SLC26A9 and NBCe1.

CFTR-mediated HCO_3_^-^secretion was not detected in F508del primary human respiratory epithelial cells. It was rescued up to ∼ 80% of the WT levelby VX-445/661/770. In contrast,TNF-α+IL-17 normalized transepithelial HCO_3_^-^transportand ASL acidic pH. This was related to anincrease in SLC26A4 and CFTR transcript levels.VX-445/661/770 induced an increase in pH only in the context of inflammation.Effects on HCO_3_^-^ transport werenot differentbetween F508del homozygous and F508del heterozygous CF airway epithelia.

Our studies show that correction of F508del-CFTRHCO_3_^-^ is not sufficient to buffer acidic ASL and that inflammation is a key regulator of HCO_3_^-^secretion in CF airways. Prediction of the response to CFTR modulators by theratyping should take into account airway inflammation.

## 1. Introduction

Airway epithelium interfaces between the internal milieu and the external environment. It is lined by a thin layer of fluid called Airway Surface Liquid (ASL) whose composition is finely tuned. There is increasing evidence that defects in ASL pH regulation are associated with various respiratory diseases, including Cystic Fibrosis (CF)(Cutting, 2015). CF is a life limiting disease caused by mutations in the cystic fibrosis transmembrane conductance regulator (*CFTR*) gene. The most frequent mutation, p.Phe508del, F508del thereafter, is carried by around 80% of people with CF (pwCF) worldwide and results in defective CFTR protein trafficking due to protein misfolding, reduced stability at the cell surface and dysfunctional channel gating (Pranke et al., 2019).

CFTR transportstwo of the most abundant and physiologically important anions: chloride (Cl-) and bicarbonate (HCO_3_^-^) into the ASL(Quinton, 2008).HCO_3_^-^plays a key role in epithelial surface homeostasis by ensuring adequate water content, controlling volume and pH of the periciliary layer, and regulating mucin biophysical properties such as hydration. All these factors are necessary for efficient muco-ciliary clearance (MCC)(Bridges, 2012; Ferrera et al., 2021; Gorrieri et al., 2016; Quinton, 2008; Tang et al., 2016).Mutations of the CFTR channel have mainly been characterized by their disruption of Cl- transport, however they also display defective HCO_3_^-^ transport which is intimately related to CF disease(Quinton, 2001). At the pulmonary level, deficient HCO_3_^-^ secretion was shown to decrease ASL hydration, reduce its pH whichfurther compromisesmucin formation, mucociliairy clearance, and impairsthe airway host defenses(Garland et al., 2013; Hoegger et al., 2014; Shah et al., 2016; Simonin et al., 2019). The addition of HCO_3_^-^ onto the CF epithelium was shown to increase ASL height and pH(Garland et al., 2013), improve mucus viscoelastic properties(Ferrera et al., 2021; Gomez et al., 2020), restore bacterial killing (Pezzulo et al., 2012), and impede the growth and biofilm formation of pulmonary bacterial pathogens(Dobay et al., 2018).

Highly effective CFTR modulator therapy (HEMT)enables partial F508del-CFTR functional restoration. Triple combination HEMT combines 2 corrector molecules, elexacaftor (VX-445) and tezacaftor (VX-661), to process misfolded F508del-CFTR protein to the cell membrane and a potentiator, ivacaftor (VX-770), to increase channel opening. Phase 3 clinical trials with Elexacaftor/Tezacaftor/Ivacaftor (ETI) have shown dramatic improvement in lung disease and a strong positive impact on quality of life for patients carrying at least one F508del-CFTR allele(Middleton et al., 2019). This involves an improvement of CFTR-dependent Cl- transport, resulting ina better hydration of airway secretions (Morrison et al., 2022). Interestingly, Rehman and collaborators showed that ETI induced an increase in pH but this was observed only in the context of inflammation with addition of TNF-α+IL-17 to the CF epithelium(Rehman et al., 2021, 2020).The failure of F508del-CFTR correction to improve pH value in the absence of inflammation was unexpected and suggested that CFTR-dependentHCO_3_^-^ secretion and its role in ASL pH control was dependent of the inflammatory status.

These studies howeverrelied on ASL pH variation, which involves both transepithelial HCO_3_^-^ and H^+^ flux. To dissect the role of CFTR on pH homoeostasis, and the potential impact of CFTR correction in the CF lung environment we characterized HCO_3_^-^ transepithelial flux and ASL pH changesin healthy and CF epithelium, and evaluated the impact of F508del-CFTR rescue in the context of inflammation.

## 3. Results

### 3.1. VX-445/661/770 increases CFTR HCO_3_^-^ flux

pH-stat experiments enabled to quantify the HCO_3_^-^ secretion rate. Reference HCO_3_^-^ secretion rates in Wild Type (WT) respiratory epithelium are shown in **Figure 1** and **Table 1Supplemental**. Basal secretion rate across WT respiratory cells (0.24± 0.04 µEqh^-1^∙cm^-2^)was increased by forskolin (Fsk)/M3-isobutyl-1-methylxanthine(IBMX)+Genistein (up to 0.51± 0.04µEqh^-1^∙cm^-2^)withsubsequent complete inhibitionby Inh-172.

**Figure 1.**
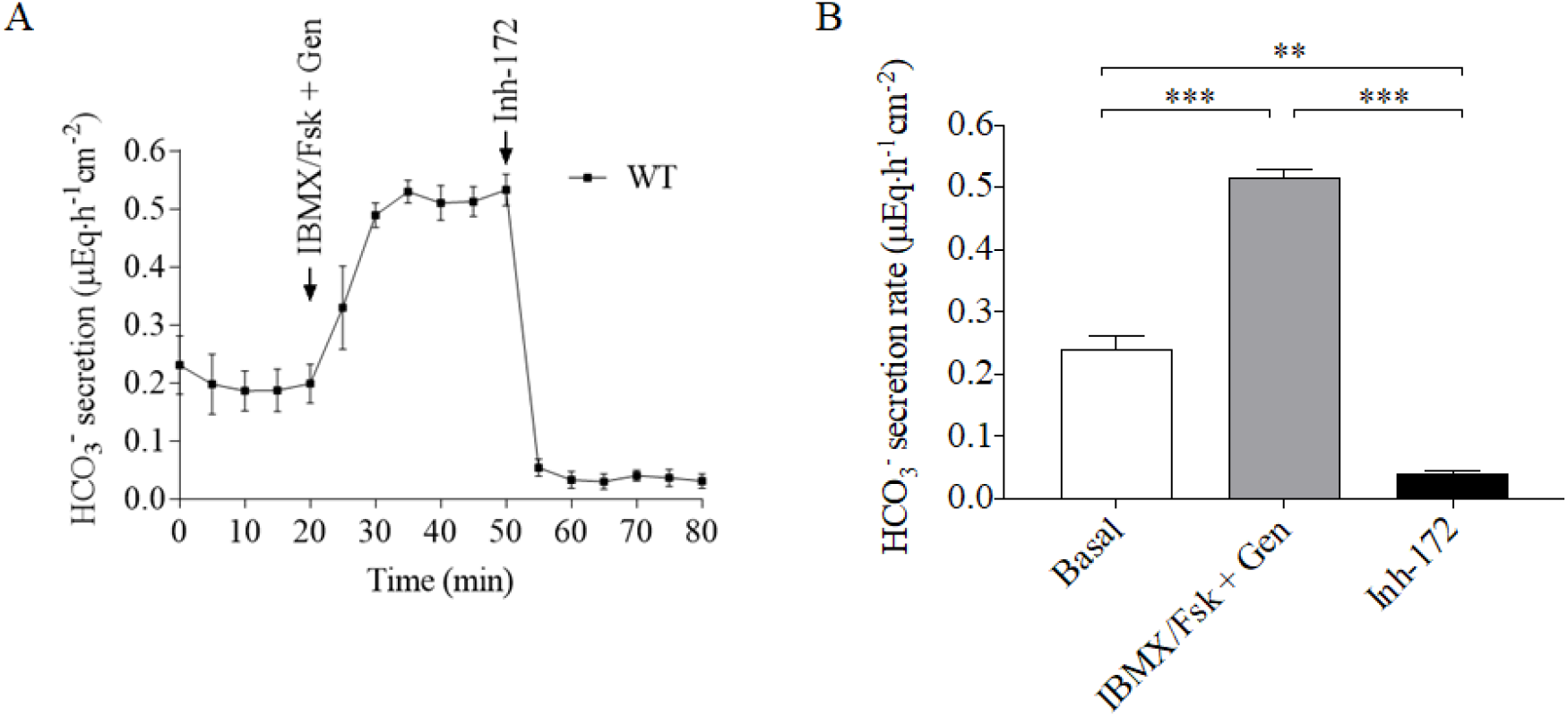
Bicarbonatesecretion in respiratory epitheliumexpressing WT CFTR. HCO_3_^-^fluxis measured in WThuman respiratory epitheliumby pH-stat in the presence of amiloride (Basal) and after successive addition of Fsk/IBMX (10µM/100 µM) + Genistein (Gen) (100 µM) andInh-172 (10 µM). (A) Representative tracing (B) Summary of 5 experiments. Results are presented as Mean ± SD (n=5). Comparison by Wilcoxon matched-pairs signed rank test.**p< 0.01; ***p< 0.001.

Representative tracings of HCO_3_^-^ fluxes in F508del human respiratory epithelial cells in DMSO and VX-445/661/770 conditions are shown in **Figure 2 Panel A**. Control DMSO treated airway epithelia displayed noHCO_3_^-^ secretion neither at basal state norafter CFTR activation by Fsk/IBMX + Genistein. In contrast, VX-445/661/770 treatment elevated basal HCO_3_^-^ secretion reaching 78±12% (0.18 ± 0.03µEqh^-1^∙cm^-2^) of the secretion level observed in WT human respiratory epithelial cells (**Figure 2, Panel A and B**). The addition of Fsk/IBMX+Genistein increased CFTR dependent HCO_3_^-^ secretionup to 70 ±29% of the WT level (**Figure 2 Panel A and C, Table 2Supplemental)**. Inh-172 completely inhibited this HCO_3_^-^ secretion, providing evidence that it was due to CFTR (**Figure 2 Panel A and D, Table 5 Supplemental)**.There was no significant difference in HCO_3_^-^ secretion rates between homozygous and heterozygous F508del cells (**Figure 2 Panel B**,**C**,**D**, and **Table 1Supplemental)**.

**Figure 2.**
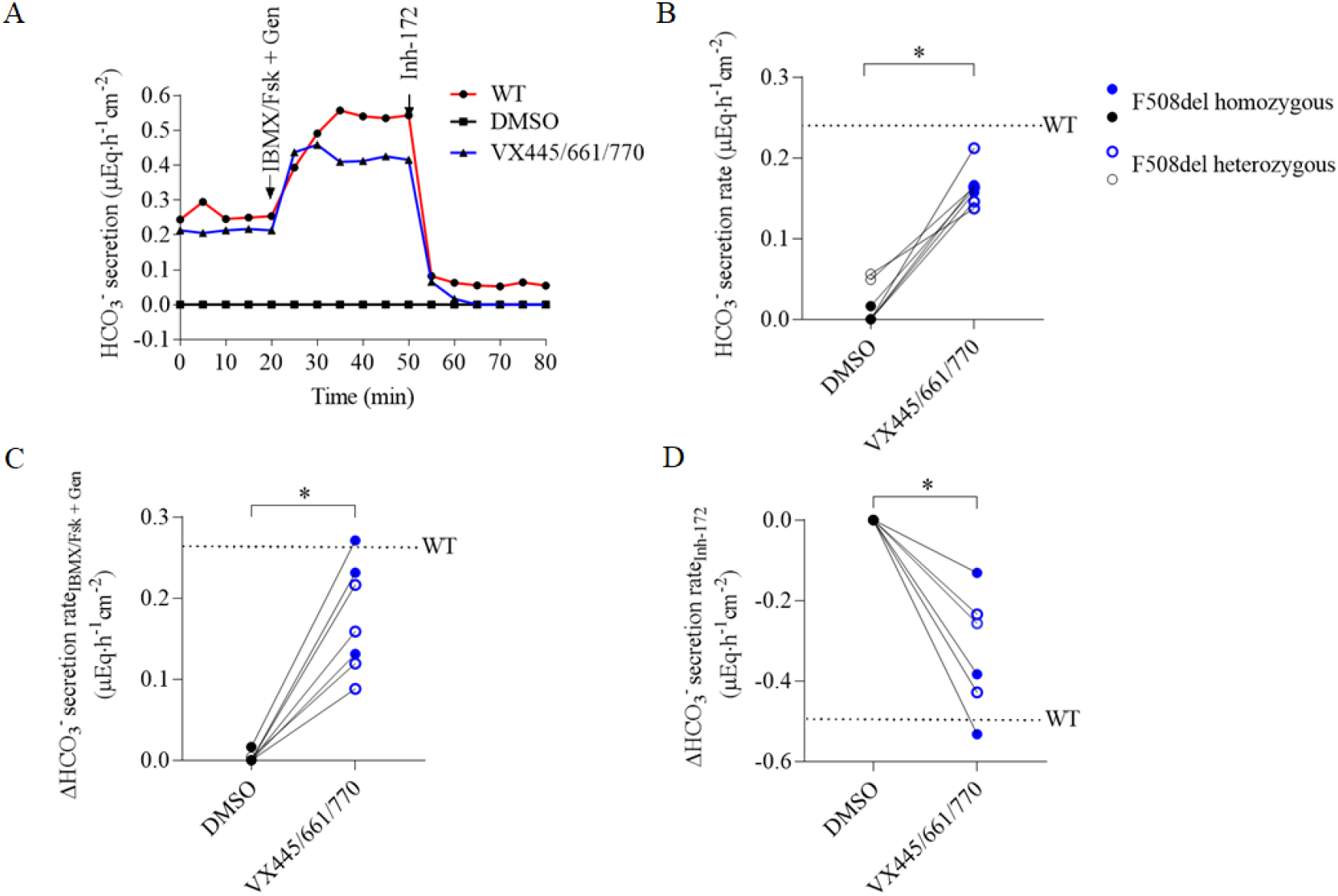
VX-445/661/770 increases F508del CFTR bicarbonate secretion in respiratory epithelial cells. HCO_3_^-^fluxis measured in WT and F508del airway epithelia by pH-stat at baseline in the presence ofamiloride (Basal) and after successive addition of Fsk/IBMX (10µM/100 µM) + Genistein(Gen)(100 µM) and Inh-172 (10 µM). Tracings were recorded in WT cells (red) and CF cells after a 48-hour incubation with DMSO (black) or VX-445(3µM)/VX-661(3µM)/VX-770 (100 nM) (blue). (A) Representative tracing of pH-stat experiment of human respiratory epithelial cells from a healthy controland a patient carrying homozygous F508del mutation. (B) Individual basal HCO_3_^-^ secretion rate changes in presence of Amiloride (100µM) between DMSO and VX-445/661/770 treated human respiratory epithelial cells(n=8). Dotted line represents the mean baseline secretion rate of WT cells. F508del homozygous human respiratory epithelial cells represented as full circles, heterozygous cells as open circles. (C) Individual HCO_3_^-^ secretion rate changes in response to Fsk/IBMX+ Genistein after a 48-hour incubation of human respiratory epithelial cells (n=7) withDMSO or VX-445/661/770. Dotted line represents the mean ofWT secretion rate change. (D) Individual HCO_3_^-^ secretion rate changes in response to Inh-172 after a 48-hour incubation of human respiratory epithelial cells (n=7) withDMSO or VX-445/661/770. Dotted line represents the mean of WT secretion rate change. Comparison by Wilcoxon matched-pairs signed rank test.* p< 0.05

### 3.2. Inflammation enhancesCFTR HCO_3_^-^ transport correction in F508delhuman respiratory epithelial cells

We next investigated the effect of inflammation alone and in combination with VX-445/661/770 on HCO_3_^-^ secretion**(Figure 3, Table 2Supplemental)**. Incubation with IL-17/TNF-*α*increased HCO_3_^-^ secretion above the WT level at basal state (comparison with WT,NS) (**Figure 3 Panel A and B, Table 2Supplemental)**.The changeafter stimulation by Fsk/IBMX + Genistein was up to 87 ± 41% of the WT (**Figure 3 Panel A and C, Table 2Supplemental)**. HCO_3_^-^ flux was partially inhibited by Inh-172, and complete inhibition was achieved by the subsequent addition of YS-01 (SLC26A4 inhibitor) and GlyH-101 (respective part in the inhibition of Inh-172, YS-01 and GlyH-101 : 37 ± 2%,; 33 ± 1.5%;29 ± 1%,)(**Figure 3 Panel A and D, Table 2Supplemental)**.

**Figure 3.**
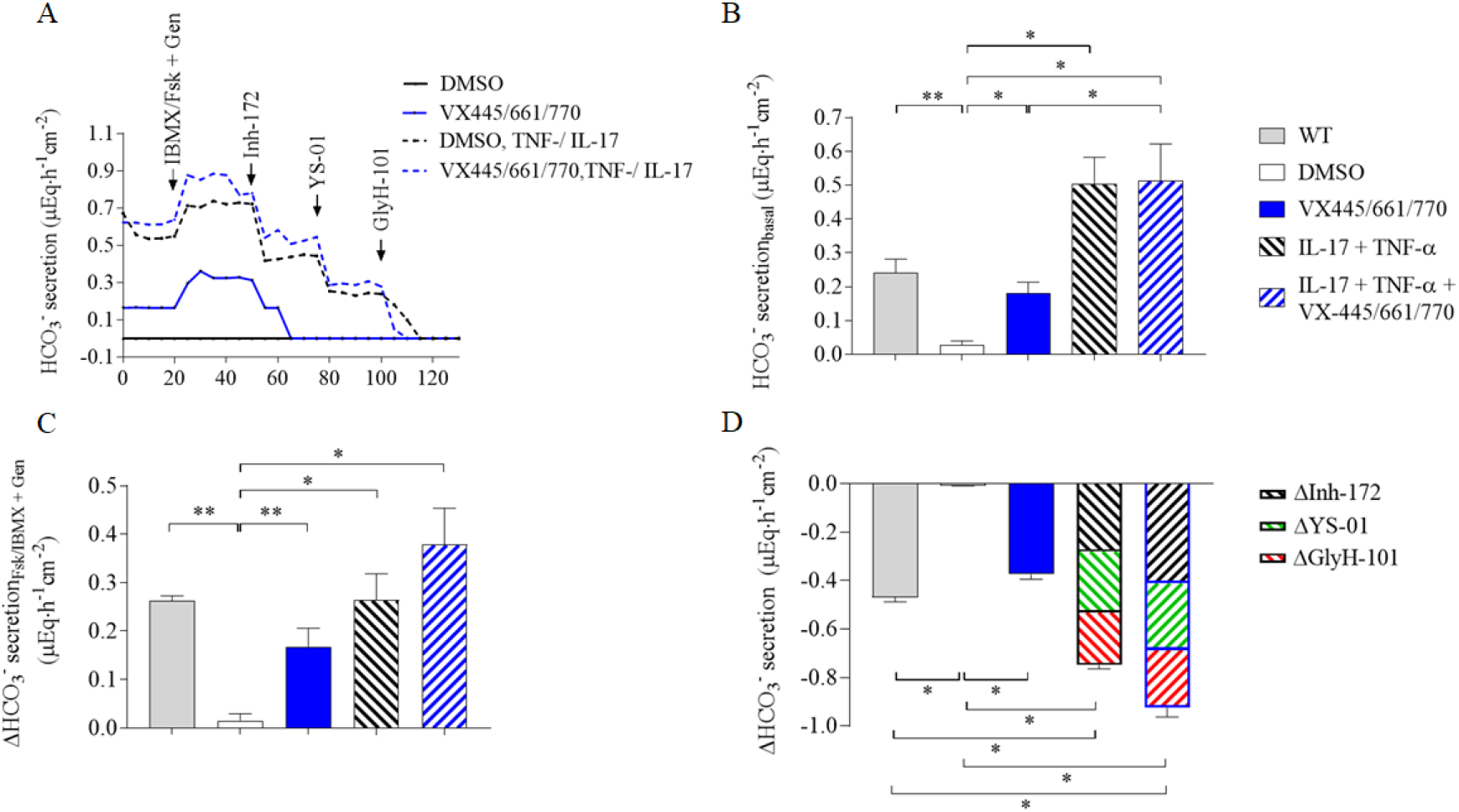
Inflammation increases bicarbonate transport in human respiratory epithelial cells. HCO_3_^-^fluxis measured in WT and F508del human respiratory epithelial cellsby pH-stat at baseline in the presence of amiloride (Basal) and after successive addition of Fsk//IBMX (10µM/100 µM) + Genistein (100 µM) and Inh-172 (10 µM), YS-01 (5µM) and GlyH-101 (10µM). CF epithelia were from 2F508del homozygous patientsand 2 F508delpatientsin *trans*with a minimal function mutation(R560K and 1717+1G>T). Tracings were recorded after a 48-hour incubation with DMSO (black line), VX-445(3µM)/VX-661(3µM)/VX-770(100 nM) (blue plain line),TNF-α(10ng/mL)/IL-17(20ng/mL) (blackdashed line) and TNF-α/IL-17 + VX-445/661/770 (bluedashed line). (A) Representative tracing of pH-stat experiment of human respiratory epithelial cells from a F508del homozygous patient. (B) BasalHCO_3_^-^ secretion inWT andCFhumanrespiratory epithelial cells after 48-hours incubation withDMSO, VX-445/661/770, TNF-α/IL-17 and TNF-α/IL-17 + VX-445/661/770. (C) HCO_3_^-^ secretion rate changesin response to Fsk/IBMX + GenisteininWT and CFhuman respiratory epithelial cells after 48-hour incubation in DMSO, VX-445/661/770, TNF-α/IL-17 and TNF-α/IL-17 + VX-445/661/770. (D) HCO_3_^-^ secretion rate changesafterFsk/IBMX + Genisteinactivation in response to Inh-172 (black lines), YS-01 (green lines) and GlyH-101 (red lines) inWT and CFhuman respiratory epithelial cells after 48-hour incubationin DMSO, VX-445/661/770, TNF-α/IL-17 and TNF-α/IL-17 + VX-445/661/770. Comparison by Mann Whiney signed rank test for WT *versus*CF cells andWilcoxon matched-pairs signed rank testfor comparisons between CF cells. p-value as follows: * p< 0.05; **p< 0.01;other comparisons were not significant

The combination of VX-445/661/770 with TNF-αand IL-17 did not modify the basal HCO_3_^-^ secretion rate orthe cAMP activated HCO_3_^-^ flux, as compared to TNF-α/IL-17 alone. In contrast, HCO_3_^-^ flux inhibition wasincreased, mainly the Inh-172 response (p<0.05) (respective part in the inhibition of Inh-172, YS-01 and GluH-101 : 45 ± 2%,; 29 ± 6%,; 26± 1.5%).

### 3.3. Inflammation but not VX-445/661/770alone induce ASL pH alkalinization

VX-445/661/770 did not significantly changeASL pH in comparison to DMSO control conditions(from 7.12(0.11) to 7.14(0.11))(**Figure 4 Panel A)**. In contrast, TNF-αand IL-17 significantly increased ASL pH to 7.37(0.04) (p=0.009)as shown in **Figure 4 Panel A**. The combination of VX-445/661/770 with TNF-α/IL-17 induced a furthersmall but significant change in ASL pH to 7.39(0.01)(p=0.03), similar to values observed in WT cells (Simonin et al, 2019).

**Figure 4.**
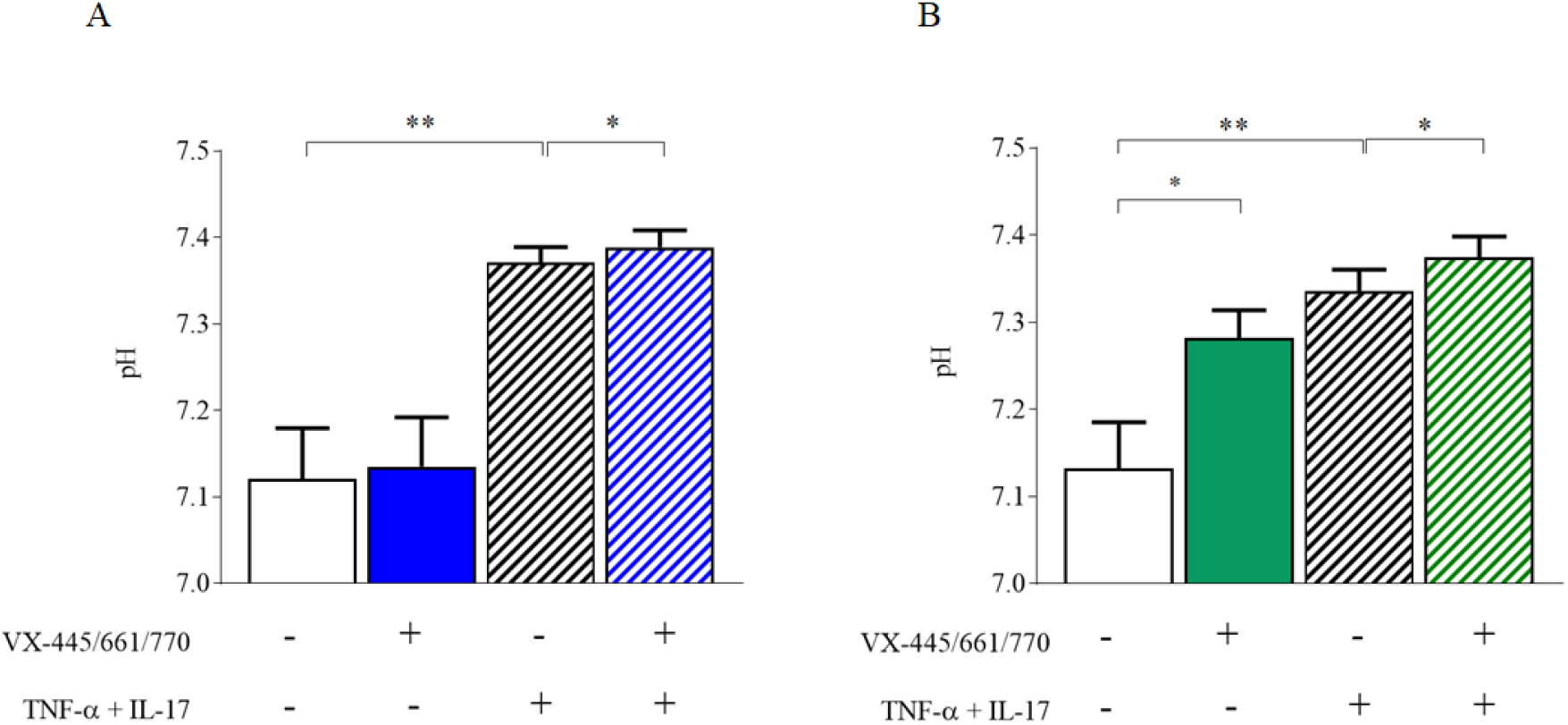
Airway surface liquid (ASL) pH of F508del human respiratory epithelial cells is abnormally acidic and is increased by VX-445/661/770 and TNF-α+IL-17 combination. ASL pH of human respiratory epithelial cellswas measured in physiological conditions (37°C, 5% CO_2_) after an 8-hour apical incubation of 50µL physiological Ringer’s solution (A) or Cl--free Ringer’s solution (B). (A) ASL pH assessed in physiological conditions in human respiratory epithelial cellsfrom F508del homozygous patients in control DMSO conditions (n=4) *versus* VX-445/661/770 (n=4); IL-17+TNF-α (n=6); IL-17+TNF-α + VX-445/661/770 (n=6). (B) ASL pH assessed in Cl--free conditions in human respiratory epithelial cells from F508del homozygous patients in control DMSO conditions (n=9) *versus*VX-445/661/770 (n=15), IL-17+TNF-α (n=9), IL-17+TNF-α + VX-445/661/770 (n=8). Data are presented as mean ± S.D.Comparison by Wilcoxon matched-pairs signed rank test p-value as follows: * p< 0.05; **p< 0.01.

To investigate the respective role of CFTR and SLC26A4 in pH homeostasis, we performed ASL pH measurementsin Cl-free solutions where SLC26A4 activity was abrogated. Underthese conditions, ASL pH of DMSO treated F508del airway cells was not significantly modified(7.12 vs 7.14 in Cl- conditions)while it significantly increasedfrom 7.14(0.11) to 7.28(0.12)when the cells were incubated withVX-445/661/770, thus unmasking a CFTR-dependent pH correction (**Figure 4 Panel B)**. In pro-inflammatory conditions, ASL pH was significantly increased up to 7.34 (0.08) (p=0.0023 *versus* DMSO but NS *versus*VX-445/661/770), with an additional effect of VX-445/661/770 reaching values similar as those obtained in physiological conditions(7.38(0.07)).

### 3.4. Inflammation increases CFTR and SLC26A4 expression

VX-445/661/770 did not modify the transcript levels of *CFTR, SLC26A4, SLC26A9* or *NBCe1*(**Figure 5**). TNF**-α/**IL-17 incubation increased SLC26A4 transcript levelsby more than 40-fold (p<0.001), but in contrast *CFTR* transcript levelsonly by 1.5-fold (p<0.01),while it did not significantly change*NBCe1* transcript levels. For *SLC26A9*, there was a trend for increase of the transcript level upon TNF-*α*/IL-17, however, this did not reach the significant level(**Figure 5**).

**Figure 5.**
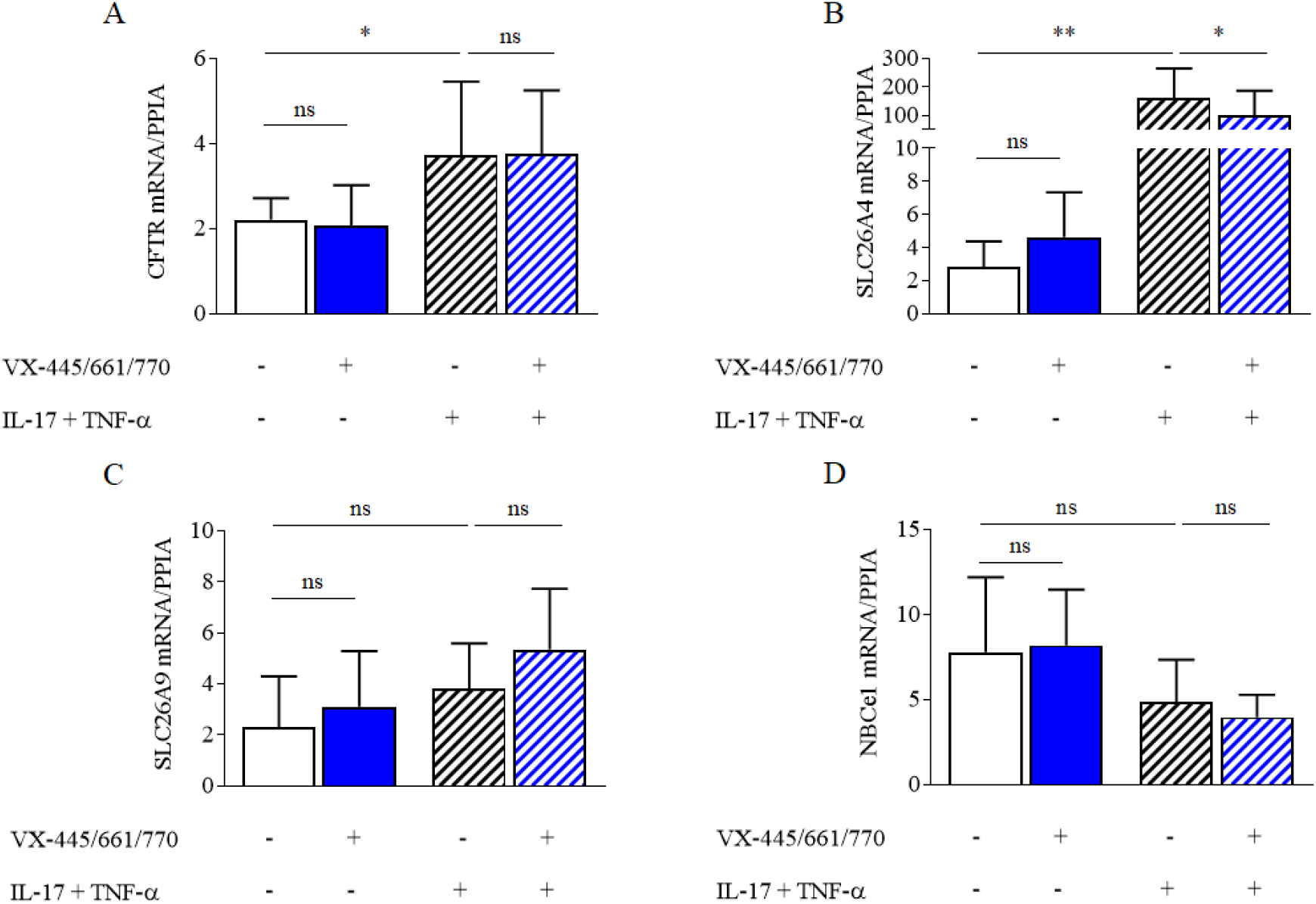
Main bases transporters expressionin primary F508del/F508del human respiratory epithelial cells by RTqPCR. Transcript expression levels of *CFTR* (A), *SLC26A4*(Pendrin) (B), *SLC26A9* (C) and *NBCe1* (D) were analysed using real-time RT–PCR in ALI differentiated primary human bronchial epithelial cells from F508del homozygous patients. Transcript levels were analysed in control DMSO condition (n=8, full white bars), VX-445/661/770 (n=4, full blue bars), IL-17+TNF-α (n=8, Stripped white bars), IL-17+TNF-α + VX-445/661/770 (n=8, stripped blue bars) and normalized with cyclophilin transcript (*PPIA*) as an internal control. When indicated, cells were treated with IL-17 (20 ng/mL) and TNF-α (10 ng/mL) and/or, VX-770 (100nM), VX-445 (3µM) and VX-661 (3µM) for 48h. Data are presented as mean ± S.D. andstatistical analysis is performed with Wilcoxon matched-pairs signed rank test(* *P*<0.05, ** *P*<0.01; ns: non significant).

## 4. Discussion

There are still few pharmacological data on rescue of HCO_3_^-^ secretion by CFTR correction in CF airway epithelium. Our results show that F508del-CFTR correction by VX-445/661/770 rescues HCO_3_^-^ transport up to 80% of the normal level in primary cultures of airway epithelium. Inflammation, e.g the TNFα/IL-17 cocktail, massively increasesHCO_3_^-^ transport, involving both CFTR and SLC26A4. CFTR correction in the context of inflammation further increased HCO_3_^-^ secretion by CFTR. Strikingly, the increase in CFTR-mediated HCO_3_^-^ secretion had a marginal impact on ASL pH, in contrast toTNF-α/IL-17 induced inflammation.

### F508del CFTR correction restores HCO3^-^ transport but not ASL pH

Similar observations were recently reported in intestinal F508del organoids where VX-445/661/770 induced an increase in CFTR-dependent HCO_3_^-^ transport (Bijvelds et al., 2022; Ciciriello et al., 2022) and in D1152H primary respiratory cells where ivacaftor improved selective HCO_3_^-^ transport defect (Laselva et al., 2020). Interestingly, in these reports as in our study, carryingF508del on both alleles did not increase the level of HCO_3_^-^ transport correction, in comparison to carrying F508del on a single allele. All these results were obtained from short circuit currents in Cl- free medium, which only enables to indirectly assess HCO_3_^-^ transport by assessing the transepithelial potential difference. Here, by using the pH-stat titration system, a powerful technique which maintains a constant pH of apical compartmentas hydrogen ions are consumed by HCO_3_^-^ secretion, we demonstrate that VX-445/661/770 treatment increases transepithelial HCO_3_^-^ fluxes. The increase of secretion in response to forskolin+genistein and its complete inhibition by Inh-172 indicates that this effect is due to CFTR. Nevertheless, despite this massive increase in CFTR-dependent HCO_3_^-^ transepithelial transport, the effect of F508del-CFTR rescue on ASL pH of cultured airway epithelia was marginal. Thisconfirmsthe observations byRehman and Morrisson(Morrison et al., 2022; Rehman et al., 2021)and provides evidence that correction of the CFTR channel defect alone is not sufficient to buffer acidic ASL of CF airways.In contrast, Ludovico et al. showedthat VX445/661/770 alkalinized the pH to WT levels(Ludovico et al., 2022). The reason of this discrepancy is unclear but may be related to differences in media culture and differential expression of HCO_3_^-^ transporters(Livnat et al., 2023).

### Functional interplay between CFTR and SLC26A4

Airway inflammation is a hallmark of CF disease observed from the first days of life, mainly driven by neutrophilic inflammation and various cytokines such as TNF-α and IL-17(Lukacs et al., 1995; Sly et al., 2013; Tan et al., 2011). TNFα/IL-17 inflammatory cytokines combination not only modulate innate anti-infective defense(McAleer and Kolls, 2011), but our data point toa massive increase of HCO_3_^-^ secretion both at basal state and after CFTR activation by cAMP (Fsk/IBMX). In addition, pH-stat experiments enabled us to dissect the participation of other transporters in ASL pH homeostasis.They clearly discriminated SLC26A4 electroneutral Cl-/HCO_3_^-^ exchange, as the complete inhibition of HCO_3_^-^ flux required the subsequent addition of YS-01 SLC26A4 inhibitor(Lee et al., 2020) and GlyH-101. This proinflammatory condition was associated with pH normalization of F508del ASL.Our results are consistent with Rehman’s observations(Rehman et al., 2020)indicating SLC26A4 as a key player in CFASL alkalinization, and the recent publication of Guidone et al. showing that YS-01 significantly decreased ASL pHin both CF and non-CF epithelia treated with IL-17/TNF-α (Guidone et al., 2022). However, in contrast to our studies, Guidone et al. observed significant ASL pH alkalinization only after stimulation ofIL-17/TNF-α treated cells by isoproterenol (Guidone et al., 2022). This discrepancy might be relatedtocell culturemedia that impact on gene expression and activity of ion channels (Saint-Criq et al., 2020). As TNF-α/IL-17 increase*SLC26A4* and *CFTR* transcript expression, one could hypothesize that the modification of pH, driven by inflammation, could be related toSLC26A4 activity, both directly by increasing HCO_3_^-^ secretion bySLC26A4 and CFTR, and indirectly via its functional coupling to corrected F508del CFTR which would fuelSLC26A4 with Cl-(Garnett et al., 2011; Kim et al., 2019). SLC26A4 appears therefore as a key player of pH homeostasis in inflammatory physiological conditions. However, the observation thatcorrection of CFTRfurther increases HCO_3_^-^ secretion by CFTRininflammatory conditions, and thatpH is significantly increased in Cl-free SLC26A4 defective conditions unravels the role ofCFTR in ASL pH homeostasis.

The fact that GlyH-101 was necessary to completely inhibit the HCO_3_^-^ transport in the context of inflammation suggests that additional HCO_3_^-^ transporters other than CFTR andSLC26A4 may be involved in this inflammation-induced alkalinization(Liu et al., 2022; Zając et al., 2021).Indeed, while Inh-172 is reported as being very specific of the CFTR gating(Caci et al., 2008),GlyH-101 occludes the CFTR pore, but is also active on Anoctamin,bestrophin (Bai et al., 2021), voltage gated channels (Lin et al., 2023)and SLC26A9(Bertrand et al., 2009; Liu et al., 2022). It it is still debated whether SLC26A9 contributes to HCO_3_^-^ secretion directly as suggested by arecent study on perfused mouse bronchioles and nasal cells(Jo et al., 2022; Liu et al., 2022)or via reciprocal regulation with CFTR, as suggested by the observation that correction of F508del-CFTR also restored SLC26A9 constitutive activity in airway cells(Bertrand et al., 2017, 2009; Larsen et al., 2021).Our experiments showthat*SLC26A9* transcripts arepresent in airways and display a trend to increase upon inflammatory trigger, however at a not significant level. This increase may be responsible for stabilization of corrected F508del-CFTR at the plasma membrane as shown by Pinto et al.(Pinto et al., 2021) and thus contribute indirectly to the higher HCO_3_^-^ fluxes mediated by CFTR observed in pH-stat experiments(Balázs and Mall, 2018).

### Physiopathological relevance of our findings

Altogether, ourresultssuggest that, in physiological conditions, SLC26A4is the main driver of pH regulation in the bronchial epithelium, CFTR intervening indirectly by fueling the exchanger with Cl-. Nevertheless, CFTR HCO_3_^-^ secretion may berequired in specific acidic situations, such as gastric acid inhalation, to efficiently and rapidly buffer the ASL pH.This has been shown in murine duodenum where low luminal pH activates transcellular HCO_3_^-^ transport that requires CFTR (Hogan et al., 1997). While a proinflammatory trigger considerably increases HCO_3_^-^ flux, itdoes not increase pH tophysiological values. Indeed, apart from pH regulation, restoration of physiological levels of HCO_3_^-^ into the airway microenvironment, by a proximal paracrine effect, would also favorrapid mucin expansion, resulting in improved mucocilary clearance and decreased airway obstruction and inflammation(Quinton, 2008). Downstream effects could also restore intrinsic immune defenses by augmenting formation of neutrophil extracellular traps by neutrophils and sentization of *P. aeruginosa* to the antimicrobial peptide cathelicidin LL-37(Siew et al., 2022).

These results suggest that a full efficiency of CFTR modulation might require a residual “background” of CFTR inflammation. This is consistent with *in vivo* studies which showed that nasal mucosa pH was abnormally acidic in CF neonates but normal from 3 months of age after airway inflammation initiation(Abou Alaiwa et al., 2014; Schultz et al., 2017) and apositive correlation between level of airway inflammation and the respiratory improvement in G551D patients treated with ivacaftor reported by Rehman et al.(Rehman et al., 2021). Several studies report that CFTR modulators abrogateairway inflammation which may then jeopardize the effect on pH(Lepissier et al., 2023). Physiopathological relevance of these findings including theirlong term remanence is still unclear(Hisert et al., 2017).

### Study limitation

This study has limitations. First, we measured ASL pH by a pH electrode and assumed that the 50 µl Ringer solution added at the surface of airway cultures for the experiment would represent diluted ASL. However, this Ringer contains 25 mM HCO_3_^-^ and could buffer small pH variations, and thus underestimate pH changes. Second, inflammation is a complex status involving different triggers, which may behave differently(Rehman and Welsh, 2023; Roesch et al., 2018). Moreover, the concentration of cytokines used in this experiment might be far above those occurring *in vivo*.Indeed, we performed a pilot study to assess inflammatory cytokines in sputum and observed that TNF-α levels were100-fold lower in patients’sputum than the concentration applied *in vitro*(Lepissier et al., 2023). Third, our experimental design only enabled to mimic acute inflammation. It could be that CFTR rescue may differentially modulate CF epithelia subjected to chronic inflammation. Finally, ASL acid-base regulation is a very complex phenomenon involvingthe coordinated activity of all ion transporting proteins,including H^+^transporting proteins present on apical and basolateral sides of the epithelium(Zając et al., 2021). The measurement of ASL pH cannot discriminate between increase in base secretion and reduction of acid secretion(Shah et al., 2016). Regulation of ATP12A proton pump by inflammation as recently reported by Guidone et al. showed that impact on H^+^ secretion needs to be considered. Most importantly, our experiments measure global luminal pH and HCO_3_^-^ transport but may underestimate local pH modification in micro areas around submucosal glands, which express CFTR at a high level and display acidic pH environment, related to CFTR defect(Garnett et al., 2016).Local correction of HCO_3_^-^ secretion may be very relevant to act on inflammation mediated mucus viscosity.

## 5. Conclusions

Our data show that CFTR correctionrestores HCO_3_^-^ secretion but at a level which is not sufficient to overcome ASL pH acidosis. Inflammationis a key regulator of HCO_3_^-^ secretion in CF airways andenhances the efficacy of CFTR modulators.Prediction of the response by theratypingshould take into account airway inflammatory phenotype and its effect on ASL pH.

## Materials and Methods

### 2.1. Patients

CF patients and healthy subjects (WT) were recruited for respiratory airway cell sampling (Clinical Trials: NCT02965326). The study was approved by the Ile de France 2 Ethics Committee, and written, informed consent was obtained from each adult and from both parents for participants below 18 (AFSSAPS (ANSM) B1005423-40, n° Eudract 2010-A00392-37; CPP IDF2: 2010-05-03-3).

### 2.2. Cell culture

Human bronchial epithelial cells were isolated from bronchial explants (1-2 bifurcation) by enzymatic digestion as previously described(Golec et al., 2022). Cells were first grown in T75cm^2^ flasks, in DMEM/F-12 cell culture medium supplemented with 10% newborn calf serum, Rho kinase inhibitor Y-27632 (10µM), Piperacillin/Tazobactam 90µg/10µg/ml, Colimycin 16µg/mL and Amphotericin B 5µg/mL, and additional growth factors. For air-liquid interface (ALI) culture 350,000 human respiratory airway expanded cells suspended in amplification medium were seeded on type IV collagen-coated porous filter with a 0.33 cm^2^ surface (6.5mm diameter, 0.4µm pore size, Corning, Tewksbury, MA). UG2% medium (DMEM/F-12, supplemented with 2% Ultroser G) containing antibiotics as above, was added to the basal side of filters. After 72 hours of liquid/liquid interface, apical media was removed and cells were cultured in ALI with UG2% basal medium changed every 48-72 hours for 3-4 weeks to establish a differentiated epithelium. Transepithelial electrical resistance (RT) of cultures was measured with a chopstick voltmeter (Millicell-ERS).

Prior functional studies, cells were incubated at the basolateral side for 48 hours with DMSO 0.03% (referred as control condition), or VX-770 (100nM), VX-445 (3µM) and VX-661 (3µM), refererred as VX-445/661/770 (Selleckchem) all dissolved in DMSO.To assess inflammation-induced responses, epithelia were treated atthe basolateral side with 10ng/ml TNF-α (R&D Systems) and 20ng/ml IL-17 (R&D Systems) for 48 hours.

### 2.3. Measurement of HCO_3_^-^fluxes

To investigate the HCO_3_^-^ transepithelial transport, HCO_3_^-^ secretion rates were monitored in Ussing chambers under open-circuit conditions by the pH-stat method. A mini pH electrode connected to Automated titration workstation, temperature sensor and burette (all from Radiometer Analytical) introduced to mucosal chamber were used to automaticallytitrate 5 mM HCl to maintain the pH of solution at 7.000 ± 0.005. TitraMaster 85 Software was used to control the rate of titration, amount of titrant added and continuous measurement of solution pH. The rate of HCO_3_^-^ secretion [µEq∙h^-1^∙cm^-2^] was calculated in 5 minute intervals by noting the amount of titrant added, its concentration and the cell surface area. HCO_3_^-^ secretion rates were calculated 20-40 minutes after addition of each activator/inhibitor and considered stable when remaining constant for at least 15 minutes. Solutions composition used for pH-stat measurements are provided in *Table 3Supplemental*. Apical solution was vigorously bubbled with pure O_2_, while basolateral solution was bubbled with carbogen (95% O_2_, 5% CO_2_). The HCO_3_^-^ secretion rates were monitored in the presence of ENaC-channel blocker Amiloride (100µM, Spectrum Chemical) (basal secretion rate) and after subsequent addition of activators or inhibitors of ion transporting proteins to determine their involvement in HCO_3_^-^ transport across epithelium. This included: cAMP agonists Forskolin (Fsk) (10µM, Sigma), M3-isobutyl-1-methylxanthine(IBMX) (100µM, Sigma-Aldrich Merck) and Genistein (10µM, Sigma-Aldrich Merck) to activate and potentiate CFTR activity; the specific CFTR inhibitor Inh-172 (10µM, Sigma-Aldrich Merck), and in case of remaining HCO_3_^-^ flux, the SLC26A4 inhibitor YS-01 (5µM, Aobious) and GlyH-101 (10µM, Sigma-Aldrich Merck). This allowed toevaluate the cAMP activated HCO_3_^-^ flux (Fsk/IBMX+Genistein) and its inhibition by Inh-172, GlyH-101 and YS-01. As CFTR may be partially activated at basal state, the change after CFTR inhibition was considered as the index of CFTR activity.

### 2.4. Measurement of ASL pH

In order to measure ASL pH under physiological conditions, we designed a system with a controlled atmosphere enclosure enabling to maintain humidity, pCO_2_ at 5%, and temperature at 37°C as previously described(Simonin et al., 2019). As ASL layer was too thin to allow direct and reproducible measurement of pH without risking disruption, pH was measured after addition of 50µL solution at the cells monolayer’s apical side. Solutions composition used for physiological condition and Cl- free condition are provided in **Table 4Supplemental**. This 50µL solution, representing diluted ASL, was harvested after 8 hours of incubation, and pH was measured immediately, directly in the controlled atmosphere enclosure with a micro-combination pH-electrode (Thermo Scientific Orion 9810BN, Illkirch, Grand Est, France). Prior to each experiment, pH microelectrode was calibrated withpH 4 and pH 7 buffers.Experimental solutions were equilibrated at pH 7.4.We previously showed that in this set-up, the measured pH value did not differby more than 0.03 pH unit from the theoretical onecalculated according to the Henderson-Hasselbalch equation(Simonin et al., 2019).

### 2.5. RT-PCR, genetic analysis

200ng of total RNA was reverse-transcribed (Thermo Fischer Scientific). Real-time quantitative PCR was performed on a LightCycler (Roche Diagnostics) with the LightCyclerFastStart DNA Master SYBR Green 1 kit (Roche Diagnostics) according to the manufacturer’s protocols, except that the reaction volume was reduced 2.5-fold. Specific primers were designed using Primer 3 (free online software, **Table 5 Supplemental**). In each run, a standard curve was obtained using serial dilution of stock cDNA prepared from the mix of the different samples total RNA. The expression of the of CFTR, SLC26A4, NBCE1and SLC26A9 transcripts was normalized to the Cyclophilin-A (PPIA) expression(mean threshold cycle for PPIA = 27.5±0.1).

### 2.6. Statistical analysis

Statistical analysis was performed using GraphPad Prism® software. Data are presented as mean ± (SD). Comparisons were performed with Wilcoxon matched-pairs signed rank test or Mann-Whitney as appropriate. Correlations were assessed with Spearman test. Significant value was retained if p<0.05.

## Data Availability

All data produced in the present study are available upon reasonable request to the authors

## Supplementary Materials

**This manuscript hassupplemantary information**

## Funding

This research was funded by Vertex Innovation Award-2017 cycle. Association ABCF, Vaincre La Mucoviscidose - RC20200502648-2020 Award, Miroslaw Zajac was financed by the Polish National Agency for Academic Exchange within Bekker Program no. BPN/BEK/2021/1/00284 and by National Science Center (NCN) Poland no. 2019/35/B/NZ1/02546.

## Conflicts of Interest

I Sermet-Gaudelus (ISG) is principal investigator of Vertex initiated studies

ISG, Luis Galietta, Gilles Crambert and Gabrielle Planelles declare academic grant funded by Vertex Innovation Award.

## Notes

### Author Declarations

The study was approved by the Ile de France 2 Ethics Committee, and written, informed consent was obtained from each adult and from both parents for participants below 18 (AFSSAPS (ANSM) B1005423-40, Eudract 2010-A00392-37; CPP IDF2: 2010-05-03-3).

