## Supplementary material for "Putting Bicarbonate on the spot. Implication for theratyping in Cystic Fibrosis": Supplmental Tables

**Table 1 Supplemental. Basal secretion and bicarbonate secretion rates changes in Human respiratory epithelial cells from healthy individuals (WT) and F508del homozygous/heterozygous patients.**

|  | **Bicarbonate transport** | | | | |
| --- | --- | --- | --- | --- | --- |
|  | WT  (n=5) | F508del/F508del  (n=4) | | F508del/other  (n=4) | |
| ΔHCO_3_-secretion rate (μEqh^-1^cm^-2^) | Mean (SD) | Mean (SD) | p-value vs WT | Mean (SD) | p-value vs WT |
| Basal secretion | 0.24(0.04) | 0.02 (0.04) | <0.001 | 0.03 (0.03) | <0.001 |
| IBMX/Fsk + Genistein | 0.27(0.03) | 0.0 (0.0) | <0.001 | 0.01 (0.03) | <0.05 |
| Inh-172 | -0.47(0.01) | 0.0 (0.0) | <0.01 | -0.02 (0.05) | <0.05 |

*Comparison by Mann-Whitney test

**Table 2 Supplemental. Basal secretion and bicarbonate secretion rates changes in Human Bronchial cells from F508del patients after correction with VX-445/661/770, incubation with TNFα and IL-17 alone and in combination with VX-445/661/770**

|  | DMSO  (n=8) | VX-445/661/770  (n=8) | | TNF-α + IL-17  (n=4) | | | TNF-α + IL-17 and VX-445/661/770  (n=4) | | | |
| --- | --- | --- | --- | --- | --- | --- | --- | --- | --- | --- |
| ΔHCO_3_-secretion rate  (μEq/h^-1^cm^-2^) | Mean  (SD) | Mean  (SD) | p-value vs DMSO* | Mean  (SD) | p-value vs DMSO* | p-value vs  VX-445/661/770* | Mean  (SD) | p-value vs  DMSO* | p-value vs  VX-661/770/445* | p-valuevs  TNF-α+IL-17* |
| Basal secretion | 0.03 (0.02) | 0.18 (0.09) | <0.05 | 0.49  (0.19) | <0.05 | NS | 0.51  (0.28) | <0.05 | NS | NS |
| IBMX/Fsk + Genistein  (n=7) | 0.02  (0.04)  (n=7) | 0.17  (0.09)  (n=7) | <0.01 | 0.27  (0.12) | <0.05 | NS | 0.38  (0.17) | <0.05 | NS | NS |
| Inh-172  (n=7) | 0.0  (0.0)  (n=7) | -0.37  (0.14)  (n=7) | <0.05 | -0.28  (0.07) | <0.05 | NS | -0.41  (0.11) | <0.01 | NS | NS |
| YS-01  (n=4) | 0.0  (0.0) | 0.0  (0.0) | NS | -0.25  (0.04) | <0.05 | <0.05 | -0.27  (0.20) | <0.05 | <0.05 | NS |
| GlyH-101  (n=4) | 0.0  (0.0) | 0.0  (0.0) | NS | -0.22  (0.03) | <0.05 | <0.05 | -0.24  (0.06) | <0.05 | <0.05 | NS |
| Total inhibition | 0.02  (0.03) | -0.37  (0.14) | <0.05 | -0.75  (0.11) | <0.05 | <0.05 | -0.92 (0.07) | <0.01 | <0.05 | <0.05 |

* Comparison by Wilcoxon matched-pairs signed rank test.

**Table 3 Supplemental. Solutions for pH-stat measurements**

| **pH-Stat experiments** | |
| --- | --- |
| Apical Ringer | Basolateral Ringer |
| NaCl 120 mM  Na-gluconate 30 mM  KCl 5 mM  CaCl_2_ 1.2 mM  MgCl_2_ 1.2mM  Mannitol 10 mM | NaCl 125 mM  NaHCO_3_25 mM  KH_2_PO_4_ 0.8 mM  K_2_HPO_4_2.1 mM  CaCl_2_ 1.2mM  MgCl_2_ 1.2mM  D-Glucose 10mM |

**Table 4 Supplemental. Ringer composition for Airway Surface Liquid pH measurements**

| **pH Measurements** | | |
| --- | --- | --- |
| **Physiological condition** | | **Chloride-free condition** |
| Apical Ringer | Basolateral Ringer | Apical and basolateral Ringer |
| NaCl 115 mM  NaHCO_3_ 23.9 mM  KCl 5.2mM  CaCl_2_ 1.0 mM  MgCl_2_ 1.0 mM  D-Glucose 10mM | Culture medium | Na isethionate 125mM  NaHCO_3_ 25mM  K_2_HPO_4_ 2.4mM  KH_2_PO_4_ 0.6mM  Ca Gluconate 3mM  Mg Gluconate 2.4mM  D-Glucose 10mM |

All solutions were equilibrated to pH 7.4.

**Table 5 Supplemental. qRT-PCR primers for acid and base transporter quantification**

| **Transporter** | **Oligo sequence** |
| --- | --- |
| Pendrin (SLC26A4) | CAGGAGAGCACTGGAGGAAA  CGAGGAATGTCACACAGCTG |
| NBCe1 (SLC4A4) | TTCACGGAACTGGATGAGCT  ACTGTGGGAGAGAAGAAGCC |
| CFTR | CATGGAATTGGAGCTCGTGG  TGACTATTGCCAGGAAGCCA |
| SLC26A9 | CTGGCCCCAGAGTCGAAATTC  CTTGAGCACCGAAATCAGGAT |
